# Seroprevalence of Chagas in urban and rural indigenous populations of the south of Gran Chaco

**DOI:** 10.1101/2021.01.11.21249585

**Authors:** Carlina Colussi, Mariana Stafuza, Marcelo Nepote, Diego Mendicino

## Abstract

**Background & objectives:** In Latin America, Chagas disease is endemic, with high prevalence in rural indigenous communities and increasing prevalence in urban areas due to migration from rural areas with active vector transmission. The objective of the work was to assess differences in the prevalence of Chagas disease in urban and rural moqoit communities of the south of Gran Chaco, Santa Fe province, Argentina.

**Methods:** A seroprevalence study was conducted in six moqoit populations. Belonging to an urban community was considered as an exposure variable, whereas seropositivity for Chagas disease was considered as an outcome variable

**Results:** The results showed that 9.26% of the 702 volunteers evaluated and 3.50% of women of childbearing potential were seropositive. The prevalence of Chagas disease in urban communities was 6.41 times higher than in rural communities.

**Interpretation & conclusions:** The seroprevalence found in the moqoit community is higher than that estimated for the general population of the same region, with greater impact in the urban area than in the rural area. The decline of vector transmission in the rural area could be related to the conservation of ancestral practices and the expansion of the agricultural model.

## Introduction

Chagas disease is a parasitic infection caused by the protozoan *Trypanosoma cruzi*. It is transmitted either through triatomine hematophagous insects (vector pathway) or congenitally, transfusionally, by organ transplantation, or by intake of contaminated food (non-vector pathways) [1].

The triatomines transmitting the infection are exclusive of Latin America, with greater presence in rural areas of the Gran Chaco eco-region. This eco-region is characterized by subtropical climate, with large thermal amplitude and high summer temperatures. It is also characterized by a xerophytic forest crossed by plain rivers, low population density, scattered precarious housing, poor socioeconomic indicators, and a large proportion of the population made up of indigenous communities [2].

In the south of Gran Chaco, in Argentina, the prevalence of Chagas disease is high in rural indigenous communities with the presence of triatomines in their homes [3], [4] and has increased in urban areas due to migration from areas with active vector transmission (Chagas disease urbanization) [5], [6], [7].

The main factors associated with Chagas disease thus include migration from an endemic area and belonging to an indigenous community [8].

The aim of this work was to assess the prevalence of Chagas disease in urban and rural moqoit communities in the south of Gran Chaco, to determine whether there is an association with Chagas disease urbanization in these groups.

## Material and Methods

### Study area

The study was conducted in indigenous communities from the south of the Gran Chaco eco-region, in Santa Fe province, Argentina, between October 2012 and September 2015. In this province, the Instituto Provincial de Aborígenes Santafesinos (Provincial Institute of the Indigenous communities of Santa Fe) has recorded a total of 46 indigenous communities [9], 13 of which live in the Gran Chaco region. Ten of these belong to the moqoit ethnic group and three to the qom ethnic group. Six of the ten moqoit communities were selected because we had had previous contacts with them. Two of these six communities live within cities (urban communities), while four of them are remote rural communities.

### Study design

The study was an observational, cross-sectional, descriptive study, carried out through convenience sampling of volunteers over one year old. For the recruitment of volunteers, both health leaders and community leaders (*cacique*s) were contacted to explain the methodology and scope of the project and to obtain their consent. Health leaders invited volunteers to participate during their home visits, whereas community leaders did so during community meetings.

Following biosecurity standards for the management of biological samples, blood samples were taken from volunteers by venous puncture, either at their homes or in community meeting rooms. Blood samples were transported refrigerated to the laboratory of the National Endemia Research Center of the School of Biochemistry and Biological Sciences of the Universidad Nacional del Litoral, Santa Fe, Argentina, for processing, and analyzed according to the Guía para la Atención del Infectado por *T. cruzi* del Ministerio de Salud de la Nación (Guidelines for the assistance to people infected by *T. cruzi* of the Argentine Ministry of Health).

### Variables

Belonging to an urban community (considering urban as those with more than 2000 inhabitants, as defined by the National Institute of Statistics and Census of Argentina [10]) was considered as an exposure variable, whereas seropositivity for Chagas disease was considered as an outcome variable. To determine seropositivity, all blood samples were analyzed by means of direct hemagglutination (Chagatest HAI Wienerlab) and recombinant ELISA (Chagatest ELISA Wienerlab). In the case of mismatch, the indirect immunofluorescence test was also performed [11]. The result was considered positive or negative according to the concordance of two tests. Age, gender and locality data were recorded.

### Statistical analysis

The data were incorporated into an Excel database and analyzed with Epi-Info® 7.2.1. For the analysis, volunteers were divided into three age groups: under 19 years old, from 19 to 50 years old, and over 50 years old. The prevalence ratio was calculated with its corresponding confidence interval, using belonging to an urban community as an exposure variable and the seropositivity for Chagas disease as an outcome variable.

### Ethical aspects

Participants were explained the benefits of the analysis and likely risks of the sampling. For volunteers between 1 and 18 years of age, consent to participate was given by a responsible adult, whereas for those over 18 years old, the consent was given by the participants themselves. The results were then given to each participant and to representative institutions of the Ministry of Health of the Province (Provincial Chagas Program and Local Health Centers) to coordinate and ensure accessibility to clinical follow-up and treatment.

The project was evaluated and approved by the Ethics and Security Advisory Committee in Research of the School of Biochemistry and Biological Sciences of the Universidad Nacional del Litoral, Santa Fe, Argentina (Act No. 03/12).

## Results

The six moqoit communities studied have an estimated population of 1543 inhabitants [12]. A total of 702 volunteers of this estimated population, i.e. 45.50%, were incorporated into the study. Of these, 32.34% (227/702) belonged to the two urban communities and 67.67% (475/702) to the four rural ones. Regarding their gender, 55.70% (391/702) were female and 44.30% (311/702) were male, with an age range of 1 to 94 years.

The seroprevalence to Chagas disease was of 9.26% (65/702), increasing from 1.94% in the volunteers under 19 years old to 54.29% in those over 50. Although no significant difference was found between sexes (p>0.05), 33.50% (131/391) of the participating women were of childbearing age (15 to 44 years of age) [13], and 18.32% of these (24/131) were seropositive for the infection. These results are summarized in Table 1.

**Table 1.** Serology for Chagas disease in urban and rural moqoit communities of the Gran Chaco of Santa Fe province, Argentina, by age range. CI: confidence interval

| Age<br>range | Sex |  |  |  | Type of community |  |  |  | Positive<br>Serology |  | Total |
| --- | --- | --- | --- | --- | --- | --- | --- | --- | --- | --- | --- |
|  | Female | %<br>(CI<br>0.05) | Male | %<br>(CI<br>0.05) | Urban | %<br>(CI<br>0.05) | Rural | %<br>(CI<br>0.05) |  |  |  |
|  |  |  |  |  |  |  |  |  | N | %<br>(CI<br>0.05) |  |
| <19<br>years | 273 | 51.90<br>(47.63-56.14) | 253 | 48.10<br>(43.86-52.37) | 127 | 24.14<br>(20.68-27.98) | 399 | 75.86<br>(72.02-79.32) | 10 | 1.90<br>(1.04-3.46) | 526 |
| 19-50<br>years | 98 | 69.50<br>(61.20-76.97) | 43 | 30.50<br>(23.03-38.80) | 70 | 49.65<br>(41.12-58.18) | 71 | 50.35<br>(41.82-58.88) | 36 | 25.53<br>(18.57-33.55) | 141 |
| >50<br>years | 20 | 57.14<br>(39.35-73.68) | 15 | 57.14<br>(26.32-60.65) | 30 | 85.71<br>(69.74-95.19) | 5 | 14.29<br>(4.81-30.26) | 19 | 54.29<br>(36.65-71.17) | 35 |
| Total | 391 | 55.0<br>(52.00-59.33) | 311 | 44.30<br>(40.67-48.00) | 227 | 32.34<br>(29.98-35.88) | 475 | 67.66<br>(64.12-71.02) | 65 | 9.26<br>(7.33-11.63) | 702 |

Belonging to an urban community and having positive serology for Chagas disease were statistically significantly associated in all age ranges. The seroprevalence of Chagas disease was 6.4 times higher in the urban population than in the rural population (Table 2).

**Table 2.** Association between serology for Chagas disease in moqoit communities and condition of living in a rural area in Gran Chaco of Santa Fe province, according to age ranges. PR: prevalence ratio CI: confidence interval

| Age range | Type of community | Serology for Chagas |  |  | PR (CI 0.05) |  |
| --- | --- | --- | --- | --- | --- | --- |
|  |  | Positive | Negative | Total |  |  |
| <19 years | Urban | 5 | 122 | 127 | 3.14 (0.92-10.68) | Chi-square 3.7208 |
|  | Rural | 5 | 394 | 399 |  |  |
| 19-50 years | Urban | 25 | 45 | 70 | 2.31 (1.23-4.32) | Chi-square 7.5806 |
|  | Rural | 11 | 60 | 71 |  |  |
| >50 years | Urban | 19 | 11 | 30 | Non-defined | Fisher's exact test 0.0135 |
|  | Rural | 0 | 5 | 5 |  |  |
| Total | Urban | 49 | 178 | 227 | 6.41 (3.73-11.02) | Chi-square 60.6710 |
|  | Rural | 16 | 459 | 475 |  |  |

## Discussion

The prevalence found in the indigenous communities studied was higher than that estimated in the total population of Santa Fe province (4%) [14],[15]. These data coincide with the increased seroprevalence found in other works in indigenous communities of Gran Chaco compared to the general population [6],[16]. Since Chagas disease is an endemic disease strongly related to socio-health conditions, the results would confirm that the indigenous peoples of this eco-region are more exposed to this neglected disease than the rest of the population.

The high seroprevalence found in women of childbearing age highlights the importance of campaigns for the detection and early treatment of congenitally infected newborns. The specific etiological treatment of women of childbearing potential has also been identified as the only effective form of prevention of congenital transmission [17], [18], [19]. In addition, in line with other studies describing the “urbanization” of the disease due to the immigration of people from endemic areas [5], [6], [7], our results showed greater seroprevalence in urban than in rural indigenous populations.

In the region, the settlement of indigenous groups is not homogeneous: rural communities inhabit either their ancestral territories or a similar habitat, preserving some of their ancestral practices [20], whereas urban ones, which have been formed more recently, are the product of migration from the lands they occupied to the periphery of the cities, because of the loss of their natural habitat, i.e. the Chaqueño forest [21]. Some studies on health-disease processes in indigenous communities have identified some Chagas disease prevention practices and have pointed out that both their daily practices and the conservation of their territories operate as protective factors against this disease [22], [23], [24]. On the other hand, the lands surrounding rural communities have undergone changes in their use and coverage due to the expansion of an agricultural model based on soybean monoculture and the use of agrochemicals [25]. In extensive crops, due to exposure to the environmental concentrations of highly used pesticides, the mortality of invertebrate communities (such as those of triatomines) has increased [26]. Thus, the lower seroprevalence of Chagas disease found in indigenous rural communities could respond to the detrimental effect of the use of agrochemicals on all living beings, including Chagas disease vectors.

## Conclusion

Given the paradigm shift in the transmission of Chagas disease occurred in recent years, from the vector to the congenital pathway and from the rural to the urban environment, it is necessary to expand public health policies, directing activities not only towards vector control in rural areas, but also to the timely diagnosis and treatment of mothers and children in both urban and rural areas. Likewise, such public policies should be accompanied by a more inclusive health system capable of ensuring accessibility to the diagnosis, treatment and control of infection, especially to the most vulnerable social groups.

## Data Availability

There are not any supplement data available

## References

[1] Pérez-Molina J & Molina I. Chagas disease. The Lancet 2018; 391(10115): 82–94. http://dx.doi.org/10.1016/S0140-6736.

[2] Benedictto M, Gómez-Valencia B, Torrella S. Structural and functional characterization of the dry forest in central Argentine Chaco. Madera y Bosques 2019; 25(2). http://dx.doi.org/1021829/myb.2521611

[3] Colussi C, Stafuza M, Denner S, Nepote M, & Mendicino D. Epidemiología de la enfermedad de Chagas en comunidades Mocovíes y Criollas en el sur del Chaco Argentino. Salud Publica Mex 2016; 58(1):3–4.

[4] Lucero R, Bruses B, Cura C, Formichelli L, Juiz N, Fernández G. et al.. Chagas’ disease in Aboriginal and Creole communities from the Gran Chaco Region of Argentina: Seroprevalence and molecular parasitological characterization. Infect Genet Evol 2016; 41: 84–92.

[5] Beloscar J, Rosillo I, Lioi S, Pituelli N, Corbera M, Turco M, et al. Migración aborigen y urbanización de la enfermedad de Chagas. Rev Fed Argent Cardiol 2007; 36: 86–87.

[6] Mendicino D, Streiger M, Del Barco M, Fabbro D, & Bizai ML. Chagasic infection and related epidemiological antecedents in a low endemicity area of Argentina. Enf Emerg 2010; 12(2): 110–114.

[7] Moscatelli G, Berenstein A, Tarlovsky A, Siniawski S, Biancardi M, Ballering G, et al. Urban Chagas disease in children and women in primary care centres in Buenos Aires, Argentina. Mem Inst Oswaldo Cruz 2015; 110(5): 644–648.

[8] Fernández M, Gaspe MS, Gürtler, R. Inequalities in the social determinants of health and Chagas disease transmission risk in indigenous and creole households in the Argentine Chaco. Parasit Vectors 2019; 12(1); 184.

[9] Ministerio de Justicia y Derechos Humanos, Gobierno de la Provincia de Santa Fe 2019. Listado de comunidades aborígenes. Available: https://www.santafe.gob.ar/index.php/web/content/view/full/117260/(subtema)/93808 (Accessed 18 April 2014)

[10] Instituto Nacional de Estadísticas y Censos de Argentina. Glosario del Censo Poblacional 2010. Available: http://www.santafe.gov.ar/index.php/web/content/download/13830/66983/file/GlosarioCensoPoblacion.pdf (Accessed 23 August 2017)

[11] Streiger M & Bovero N. Indirect immunofluorescence reaction for the diagnosis of Chagas disease. Preservation of the imprints. Medicina (B Aires) 2010; 40(1):250–1.

[12] Instituto Nacional de Estadística y Censos. Encuesta Complementaria de Pueblos Indígenas (2004-2005). Available: http//www.indec.mecon.ar/micro_citios/webcenso/ECPI/pueblos/ampliada_index.asp?mode=09 (Accessed 24 March 2016)

[13] Organización Mundial de la Salud. Centro de prensa. Salud de la Mujer. 2013. Available: http://www.who.int/mediacentre/factsheets/fs334/es/ (Accessed 23 August 2017)

[14] Spillmann C, Burrone S, & Coto H. Análisis de la situación epidemiológica de la enfermedad de Chagas en Argentina: avances en el control, 2012. Rev Argent Salud Publica 2013; 40–44.

[15] Mendicino D, Colussi C, Stafuza M, Manattini S, Montemagiore S & Nepote M. Seroprevalencia de Chagas en mayores de 14 años de áreas rurales del Chaco Santafesino. Rev Fac Cien Med Univ Nac Cordoba 2019; 76(1): 47–51.

[16] Moretti E, Castro I, Franceschi C & Basso B. Chagas disease: serological and electrocardiographic studies in Wichi and Creole communities of Misión Nueva Pompeya, Chaco, Argentina. Mem Inst Oswaldo Cruz 2010; 105(5):621–626.

[17] Fabbro D, Danesi E, Olivera V, Codebó M, Denner S, Heredia C, et al. Trypanocide treatment of women infected with Trypanosoma cruzi and its effect on preventing congenital Chagas. PLoS Negl Trop Diseases 2014; 8(11).

[18] Moscatelli G, Moroni S, García-Bournissen F, Ballering G, Bisio M, Freilij H, et al. Prevention of congenital Chagas through treatment of girls and women of childbearing age. Mem Inst Oswaldo Cruz 2015; 110(4), 507–509.

[19] Murcia L, Simón M, Carrilero B, Roig M, & Segovia M. Treatment of infected women of childbearing age prevents congenital Trypanosoma cruzi infection by eliminating the parasitemia detected by PCR. J Infect Dis 2017; 215(9): 1452–1458.

[20] Cuneo F, Méndez MF, Sponton G, & Mendicino D. Conservación de las formas de alimentación ancestrales en comunidades moqoit del Chaco Argentino. Diferencias urbano rural. Spanish journal of community nutrition. 2019; 25(3): 6.

[21] Weiss L, Engelman J, Valverde S. Pueblos indígenas urbanos en Argentina: estado de la cuestión. Revista Pilquen 2013;16(1):1–14.

[22] Scarpa G, & Rosso C. La etnobotánica moqoit inédita de Raúl Martínez Crovetto I. Descripción, actualización y análisis de la nomenclatura indígena. Boletín de la Sociedad Argentina de Botánica 2014; 49(4): 623–647.

[23] Forsyth C. From Lemongrass to Ivermectin: Ethnomedical Management of Chagas Disease in Tropical Bolivia. Med Anthropol 2018; 37(3): 236–252.

[24] Suarez M. Medicines in the forest: Ethnobotany of wild medicinal plants in the pharmacopeia of the wichí people of Salta province (Argentina). J Ethnopharmacol 2019; 231: 525–544.

[25] Moreno M, Hoyos L, Cabido M, Catalá S & Gorla D. Exploring the association between Trypanosoma cruzi infection in rural communities and environmental changes in the southern Gran Chaco. Mem Inst Oswaldo Cruz 2012; 107(2): 231–237.

[26] Ronco A. Algunas respuestas sobre los impactos del uso de plaguicidas para el control de plagas en agroecosistemas de la Región Pampeana. Ciencia e Investigación 2015; 65(2): 63–71.

